# The incidence and in-hospital mortality of COVID-19 patients post-vaccination in eastern India

**DOI:** 10.1101/2021.07.15.21260265

**Authors:** Abhraneel Guha, Aritra Chakrabarti, Subhrojyoti Bhowmick, Saibal Das, Rahul Khandelwal, Aditya Kumar, Ajoy Krishna Sarkar, Anupam Das, Krishnangshu Ray, Sujit KarPurkayastha

## Abstract

**Objectives:** The comparable effectiveness of Covishield and Covaxin vaccines has not been studied. We compared the effectiveness of Covishield and Covaxin vaccines against moderate to severe COVID-19.

**Methods:** In this retrospective observational study, we collected data of patients who were admitted with moderate to severe COVID-19. The vaccination status and comorbidities of the patients were documented. The incidence and in-hospital mortality of COVID-19 patients was assessed. Univariate analysis was performed to determine the risk factors of in-hospital mortality.

**Results:** Of 294 patients, 5.1% (n=15) received Covaxin and 26.5% (n=78) received Covishield; 68.4% (n=201) patients were unvaccinated. Of patients who were vaccinated and contracted COVID-19, 24.8% (n=73) had taken the first dose and 6.8% (n=20) had taken the second dose of either vaccine. The in-hospital mortality rate was 13.6% (n=40). 24/40 (60%) people who had hospital mortality were unvaccinated.3/40(7.5%) had succumbed to death after receiving double dose of Covishield, 11/40 (27.5%) had succumbed to death after receiving single dose of Covishield, 2/40(5%) had succumbed to death after receiving single dose of Covaxin, none had reported infection after receiving second dose of Covaxin. No significant association was found with the type of vaccine and the in-hospital mortality (p=0.23). Significant associations with in-hospital mortality were found with the interval before COVID-19 disease and vaccination (OR, 3.02; p=0.01); and the presence of diabetes mellitus (OR, 2.13; p=0.02), cardiovascular diseases (OR, 2.11; p<0.001), and malignancy (OR: 2.33; p=0.0325).

**Conclusion:** There was no significant difference in the effectiveness of Covaxin and Covishield in terms of the incidence of COVID-19 and in-hospital mortality. Diabetes mellitus, cardiovascular diseases, and malignancies had a significant association with in-hospital mortality in patients with moderate to severe COVID-19.

## Introduction

COVID-19 causes a variety of symptoms like fever, dry cough, dyspnea, fatigue, body ache, headache, abdominal pain, diarrhea.^[1]^ Several vaccines have been tried to prevent COVID-The efficacy of the Oxford-AstraZeneca Vaccine (Covishield™) for the pre-specified primary analysis against the primary endpoint of COVID-19 occurring more than 14 days after the second dose was 70.4%. Surprisingly, however, the efficacy was substantially lower in the SD/SD (2 standard doses) than in the LD/SD (low dose/standard dose) which remained after accounting for differences in age and time between doses.^[2]^

On the other hand the Bharat Biotech vaccine Covaxin™ was found to be a safe and tolerable vaccine with minimal and minor adverse effect profile.^[3]^ In phase II clinical trial of Covaxin™, following two-dose immunization schedule at days 0 and 28 with 6 and 3 μg antigen with imidazoquinoline, the vaccine candidate showed noteworthy results in plaque reduction neutralization test based assay, with the seroconversion rates of neutralizing antibodies being 98.6%.^[4]^

While both vaccines elicited immune response, seropositivity rates to anti-spike antibody were significantly higher in Covishield™ recipients compared to Covaxin™ after the first dose.^[5]^ However, the comparative effectiveness of these two vaccines has not been studied much in India. We compared the effectiveness of Covishield™ and vaccines Covaxin™ against moderate to severe COVID-19.

## Materials and Methods

This study was performed after obtaining approval from the institutional ethics committee. In this retrospective observational study, we collected data of patients who were admitted with moderate to severe COVID-19 from 15 March to 31 May 2021 at Peerless Hospital & B K Roy Research Center in Kolkata, India from medical records. Mild COVID-19 patients were excluded. The demographic characteristics of all patients were noted. Details of their vaccination status and comorbidities were documented.

The proportion of patients contracting severe COVID-19 post-vaccination and the in-hospital mortality rate were analysed. The effectiveness of Covishield™ and Covaxin™ were compard in terms of the incidence of COVID-19 and in-hospital mortality.

Descriptive statistics were used for demographic variables. The chi-square test was used to compare the categorical variables. Univariate analysis was performed to determine the risk factors of in-hospital mortality post-vaccination. All analyses were performed in SPSS version 23 (IBM, NY). A p-value of <0.05 was considered significant.

## Results

Out of 294 patients whose data were analysed, 5.1% (n=15) took Covaxin™ and 26.5% (n=78) took Covishield™; 68.4% (n=201) patients were unvaccinated. Of those who were vaccinated and contracted COVID-19, 24.8% (n=73) had taken first dose and 6.8% (n=20) had taken second dose of vaccine; 13.7% (n=32) were unable to recollect the date and dose of vaccine.24/40 (60%) people who had hospital mortality were unvaccinated.3/40(7.5%) had succumbed to death after receiving double dose of Covishield, 11/40 (27.5%) had succumbed to death after receiving single dose of Covishield, 2/40(5%) had succumbed to death after receiving single dose of Covaxin, none had reported infection after receiving second dose of Covaxin. Of various comorbidities, 34.4% (n=101) of the patients were having diabetes mellitus, 39.1% (n=115) had hypertension, 6.1% (n=18) had chronic kidney disease, 8.5% (n=18) had cardiovascular diseases, 6.8% (n=20) had lung diseases, and 1.4% (n=4) had malignancies.

The total in-hospital mortality rate was 13.6% (n=40). No significant association was found between the type of vaccine and the in-hospital mortality (p=0.23). Significant associations with in-hospital mortality were found with the interval before COVID-19 disease and vaccination (OR, 3.02; p=0.01); and the presence of diabetes mellitus (OR, 2.13; p=0.02), cardiovascular diseases (OR, 2.11; p<0.001), and malignancy (OR: 2.33; p=0.03). However, no association with in-hospital mortality was found with hypertension (OR, 0.21; p=0.63), chronic kidney disease (OR, 0.55; p=0.07), and lung diseases (OR, 0.74; p=0.85) (Table 1).

**Table 1:**
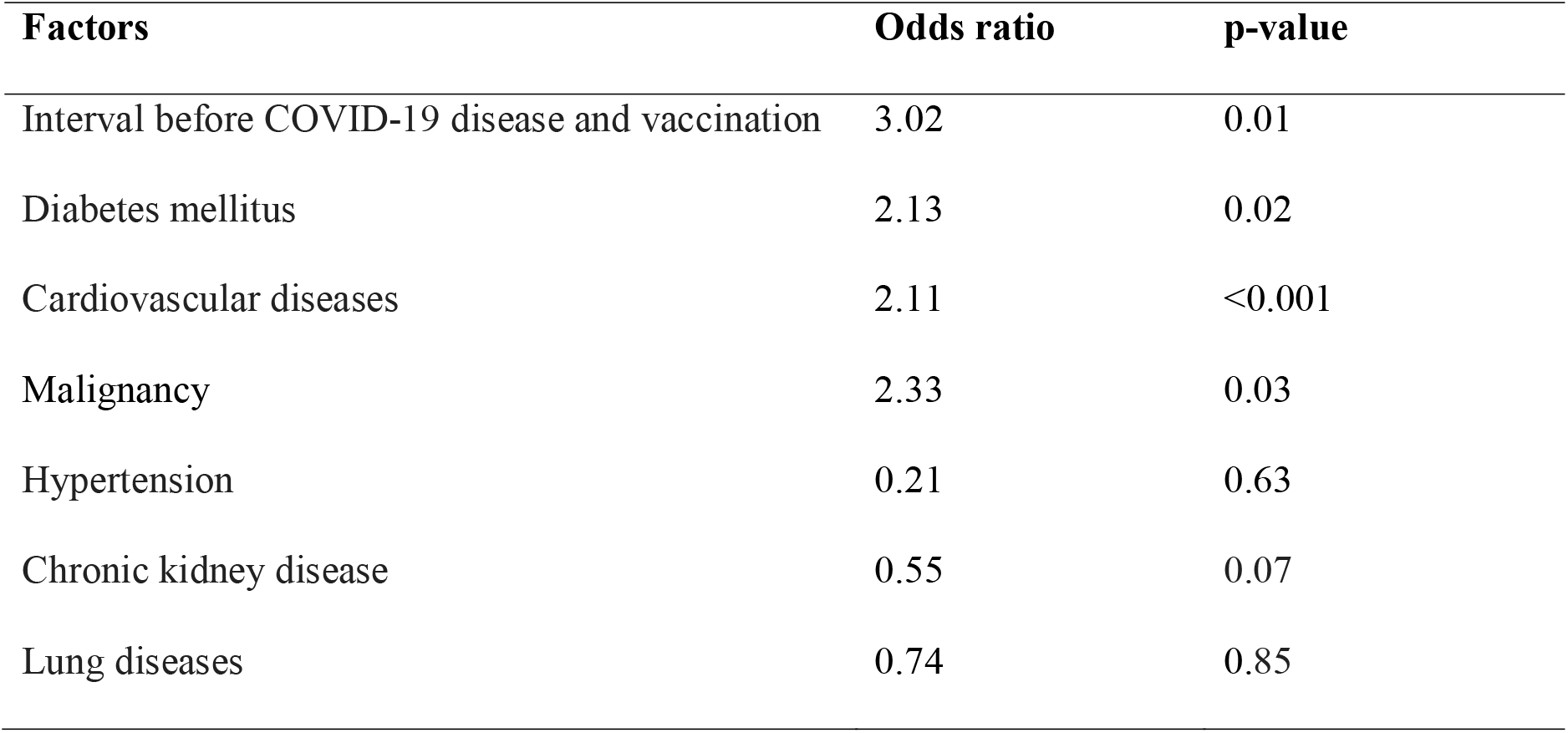
Risk factors for in-hospital mortality following complete vaccination (n=294).

## Discussion

In this study, we found that the interval between the day of COVID-19 positivity and vaccination was having a significant association with in-hospital mortality. There was no significant difference between the two vaccines. Diabetes mellitus, cardiovascular diseases, and malignancies had a significant association with in-hospital mortality in moderate to severe COVID-19.

Foy et al. concluded that prioritizing older age groups for vaccination could reduce the mortality rate.^[6]^ Iacobucci et al. suggested that a single vaccine dose was only 33% effective against the B.1.617.2 variant first detected in India. Covishield™ was 60% effective against B.1.617.2 at 2 weeks after the second dose.^[7]^ Jacob et al. did genome analysis of Indian isolates indicates that pangolin lineage B.1/B.1.1/B.1.36 (previously A2a) with D614G mutation being dominant with a possibility of more efficient transmission. The second common lineage B.6 (previously A3/A3i)^7^ in India has less transmission ability, it could be due to the lack of mutation in the spike protein. As the mutation frequency in S protein is comparatively lesser, the vaccines were expected to have a wide coverage worldwide including India.^[8]^

We found that patients who received the second dose of vaccine were better protected. Only 6.8% were infected after the second dose. The interval post-vaccination was also important. With time after the second dose, fewer patients are infected. New vaccines are on the way that can aid in the production of antibodies even against mutants.^[9]^ According to Pal et al. patients with diabetes mellitus have a poor prognosis if they are infected with COVID-19, they also suggested that patients with diabetes mellitus, cardiovascular diseases, and malignancies should be prioritized for vaccination.^[10]^

The limitations of our study include the retrospective design and the small sample size. Nonetheless, this is the first study from eastern India demonstrating the comparative effectiveness of Covaxin™ and Covishield™.

## Conclusion

The interval between vaccination and the day of COVID-19 positivity was having a significant association with in-hospital mortality. There was no significant difference in the effectiveness of Covaxin™ and Covishield™. Diabetes mellitus, cardiovascular diseases, and malignancies had a significant association with in-hospital mortality in moderate to severe COVID-19. Large-scale multicentric studies are needed to further verify our findings.

## Data Availability

All data are authentic.Data were collected and analysis was done accordingly.

